# Assessing the approach to perinatal mental health screening and treatment in maternal-child health clinics in Western Kenya

**DOI:** 10.1101/2025.08.28.25334629

**Authors:** Asterico Neema, Anna Larsen, Agnes Karingo Karume, Anne Kaggiah, Anjuli D Wagner, Barbra Richardson, Bryan Weiner, Catherine Maina, Clinton Dambuki, David Owaga, Nancy Ngumbau, Lincoln Pothan, Yuwei Wang, Alexandra Rose, Linnet Ongeri, Amritha Bhat, Keshet Ronen, John Kinuthia

## Abstract

**Background:** Integrating mental health care into well-attended health services, such as maternal-child health (MCH), offers a promising approach to increasing access to mental health care. To inform integration, we assessed the current approach to screening and treatment of perinatal mood and anxiety disorders (PMAD) in MCH facilities in Western Kenya.

**Methods:** We conducted a cross-sectional survey among the facility managers (“in-charges”) at 20 MCH facilities in Western Kenya. Trained data collectors administered questionnaires to assess facility infrastructure and human resources, perinatal mental health screening, psychological interventions, psychiatric medication availability, and level of perinatal mental health integration into standard MCH services.

**Results:** Across 20 MCH facilities, the majority were located in rural areas (16,80%), 2 peri-urban (10%), and 2 urban (10%). Facilities had a median of 37.5 medical staff (IQR: 31.0, 45.5); the most common cadre was nurses (median: 9.0, IQR: 6.0, 12.5). Under half of the facilities (8, 40%) screened for PMAD using a validated tool and documented results; most (19, 95%) reported diagnosing PMAD, yet only half (10, 53%) documented the diagnosis. The most common psychotherapy reported across the sites was supportive counseling (15, 75%). Some facilities offered evidence-based psychotherapies (e.g., cognitive behavioral therapy, problem-solving therapy, etc.), but did not report on training, supervision or guidance from the evidence-based intervention manual. Across all facilities, the availability of mental health medications was limited. Only (12, 60%) had any antidepressant available, (16, 80%) had antiepileptics, (9, 45%) had antipsychotics, and (2, 10%) had mood stabilizers available.

**Conclusion:** Mental health services in MCH clinics in Western Kenya are currently not offered in a structured and systematic manner to effectively alleviate PMAD. Available infrastructure and human resources offer an opportunity to integrate an evidence-based treatment model to improve perinatal mental health in the MCH clinics in Western Kenya.

## 1. INTRODUCTION

Perinatal mood and anxiety disorders (PMAD) are among the most common complications that occur during pregnancy or in the first year after delivery (1). PMAD is a public health problem, particularly in low- and middle-income countries (LMIC) (2). Women in LMICs have an elevated risk of PMAD compared to those in higher-income settings (3). Recent systematic reviews found that 1 in 4 perinatal women in LMICs experience depression, and 1 in 5 experience anxiety, compared to 10-15% and 7-15% prevalence, respectively, in high-income countries (4–6). A large prospective analysis in Kenya reported high prevalence (33%) of moderate-to-severe depressive symptoms among perinatal Kenyan women in public sector MCH care (7). Disproportionately high PMAD in LMICs may be associated with exposure to stressors and risk factors like a higher likelihood of socio-economic deprivation, more severe and more chronic poverty, higher co-morbidity with chronic medical conditions, sociocultural factors such as gender inequity, and poorer levels of education among women, and persisting stigma around mental health disorders(8–10). Further, mental illnesses during the perinatal period are underestimated, underassessed, and underdiagnosed. Consequently, perinatal mental health conditions are often untreated (11). Perinatal mental disorders are highly treatable when identified early yet remain leading contributors to pregnancy and postpartum-related morbidity and mortality (12). Untreated PMAD causes significant disability among perinatal women (13,14), and are associated with adverse outcomes such as preterm birth, low birth weight, insecure infant attachment, and later childhood mental health and behavioral problems (15)

Maternal child health services are well-attended in Kenya (>90%) (KDHS 2022), offering an opportunity to provide mental health services integrated into routinely attended health visits throughout pregnancy and postpartum. Integrating mental health services into commonly attended public healthcare represents a particularly relevant health system strengthening strategy in LMICs (16). Maternal child health services offer an appropriate platform for ensuring equitable, accessible, and holistic mental health services are offered to pregnant and postpartum mothers (17). To expand access to perinatal mental health, the Kenya Mental Health Action Plan 2021-2025 advocates for integrating mental health care into well-attended health services such as MCH (Health, 2015)

Effective service integration requires a comprehensive understanding of the healthcare infrastructure, providers, standard procedures, and resources currently available. However, limited information is available to guide effective perinatal mental health service integration in Kenya. To address this gap, we assessed the current approach to perinatal mental health screening and treatment in maternal and child health clinics in Western Kenya and identified opportunities for leveraging current infrastructure to expand perinatal mental health services.

## 2. METHODS

### 2.1 Study Design, population, and setting

We conducted a cross-sectional survey among the facility managers (“in-charge”) at 20 MCH facilities in Homabay, Kisumu, and Siaya counties of Western Kenya from January 30, 2024 to April 17, 2024. Overall, 47 facilities were evaluated for potential inclusion in the IPMH study, 20 health facilities were selected based on ANC volume (≥30 new ANC clients per month). Selected facilities included a range of characteristics such as rural/urban location, mental healthcare model, and facility level 3 and level 4 (“level” is a marker of health service provision hierarchy as stipulated in the Kenya Essential Package for Health (KEPH). Facilities that had active research studies evaluating perinatal mental health interventions and level 5 facilities with mental health specialists were excluded. All facility managers in the 20 health facilities who were approached to participate in the cross-sectional survey gave oral consent and responded to the facility checklist questionnaire. The consent process was documented in the researcher’s field notes, including the date, and confirmation that all required information was provided and understood, and that verbal consent was given.

### 2.2 Procedures

Trained data collectors administered tablet-based questionnaires to facility managers at each of the 20 MCH facilities.

To evaluate the availability of specific perinatal mental health services, we used the WHO Special Initiative for Mental Health Facility Checklist (WHO/UCN/MSD/ODM/2023.01 2023), which our study team adapted for the Western Kenyan context. Items in this checklist identify whether specific services (e.g., screening for mental health symptoms using a validated scale, documentation of mental health diagnostic results in medical records, prescription of psychiatric medication, etc.) are offered in a facility (yes/no). The questionnaire also evaluated which cadres of healthcare workers (e.g., obstetricians, social workers, etc.) provide each service. In the Kenyan context, the role of “medical officer” describes a provider who has completed undergraduate medical school and has not yet completed specialist training. The role of “clinical officer” describes a licensed provider with generalist clinical training.

We explored psychiatric medication availability within the facility, asking facility managers to list the top three psychopharmaceuticals most frequently prescribed in each category: 1. We explored psychiatric medication availability within the facility, asking facility managers to list the top three psychopharmaceuticals most frequently prescribed in each category: 1. Antipsychotics, 2. Antidepressants, 3. Anxiolytics, 4. Mood stabilizers, 5. Antiepileptics. Study nurses evaluated the availability of each drug specified on the day of data collection as “available today and not expired”, “available but expired”, “reported available but not seen”, or “not available today”.

Information about psychological treatments available at the facilities was elicited by listing types of interventions (e.g., problem-solving therapy, Problem Management Plus (PM+), cognitive behavioral therapy, etc.) and asking facility managers to indicate whether these are offered, by which cadres, and whether the target patient population includes perinatal women. If facility managers endorsed that a psychological therapy type was available, collectors further probed whether it was guided by a published manual, whether the published manual was reported available, and whether the data collectors observed the manual.

To understand how perinatal mental health services may differ between perinatal women living with HIV versus those without HIV, we asked whether differences existed (yes/no). In cases where differences were identified, facility managers provided qualitative descriptions of the differences. Understanding the differences helped identify gaps and inequalities in perinatal mental health service provision.

Data collectors guided facility managers in determining the “Level” of integration in that facility for perinatal mental health services within routine MCH services using the “Substance Abuse and Mental Health Services Administration (SAMHSA) Framework for Integration”. (18) The levels were: 1. Minimal collaboration, 2. Basic collaboration at a distance, 3. Basic collaboration onsite, 4. Close collaboration in a partly integrated system, 5. Close collaboration in a fully integrated system. In multiple questionnaire modules, facility managers were offered the opportunity to provide more detail as qualitative “free text” descriptions.

### 2.3 Data analysis

We used descriptive statistics (counts and proportions for binary and categorical variables, medians and interquartile ranges for continuous variables) to summarize perinatal mental health services offered within facilities based on the aspects assessed in the facility checklist. We synthesized these data to describe current availability and gaps in perinatal mental health service provision, the similarities and differences in perinatal mental health care provided to HIV-negative women vs. women living with HIV, and to evaluate the integration level of mental health services in MCH/PMTCT facilities. Statistical analyses were performed in Stata 18.0.

### 2.4 Ethical considerations

All the participants gave verbal consent to participate in the survey. This study was approved by the ethical review committees both at Kenyatta National Hospital / University of Nairobi (ERC number P425/04/2023) and at the University of Washington (IRB number 00017933).

### 2.5 Role of funding sources

The funding agency had no role in writing this manuscript or submitting it for publication.

## 3. RESULTS

### 3.1 Characteristics of Maternal Child Health Facilities Assessed in Western Kenya

Characteristics of the 20 sampled facilities are presented in Table 1. The facilities included 16 rural facilities (80%), two peri-urban facilities (10%), and two urban facilities (10%). Half of the facilities were located in Siaya county (n=10), followed by Homabay county (n=6, 30%), and Kisumu County (n=4, 20%). Over half of the facilities were sub-county hospitals (n=12, 60%) and eight (40%) were health centers. Eight facilities (40%) reported a monthly antenatal care (ANC) new patient volume of between 30-40 patients, while six (30%) had between 51-60 new ANC clients per month, four (20%) had between 41-50, and two (10%) had between 61-70 new ANC patients per month.

**Table 1.** Characteristics of Maternal Child Health Facilities Assessed in Western Kenya.

| **Facility characteristics** | **N (%) or Median (Interquartile range)** |
| --- | --- |
| County |  |
| Homa Bay | 6 (30%) |
| Kisumu | 4 (20%) |
| Siaya | 10 (50%) |
| Location category |  |
| Rural | 16 (80%) |
| Peri-urban | 2 (10%) |
| Urban | 2 (10%) |
| Health system level |  |
| Level 3 | 8 (40%) |
| Level 4 | 12 (60%) |
| Monthly antenatal care new patient volume | 43.38 (35.25-51.63) |

### 3.2 Perinatal mental health care cascade in MCH facilities in Western Kenya

Figure 1 is a representation of perinatal mental health services offered in the MCH facilities. Out of the 20 sites assessed, eight (n=8/20, 40%) reported screening for PMAD using a validated scale, of which all eight reported documenting PMAD screening results. In the 8 facilities that reported screening using validated scales, healthcare worker cadres conducting screening were most commonly nurses (n=8/8, 100%), followed by clinical officers (n=7/8, 88%). Nearly all facilities (n=19/20, 95%) reported routinely conducting assessments for PMAD diagnosis, among which only half (n=10/19, 53%) reported documenting the diagnosis. Among the 19 facilities diagnosing mental health conditions, this task was primarily conducted by clinical officers (n=17/19, 89%) followed by nurses (n=13/19, 68%).

**Figure 1.**
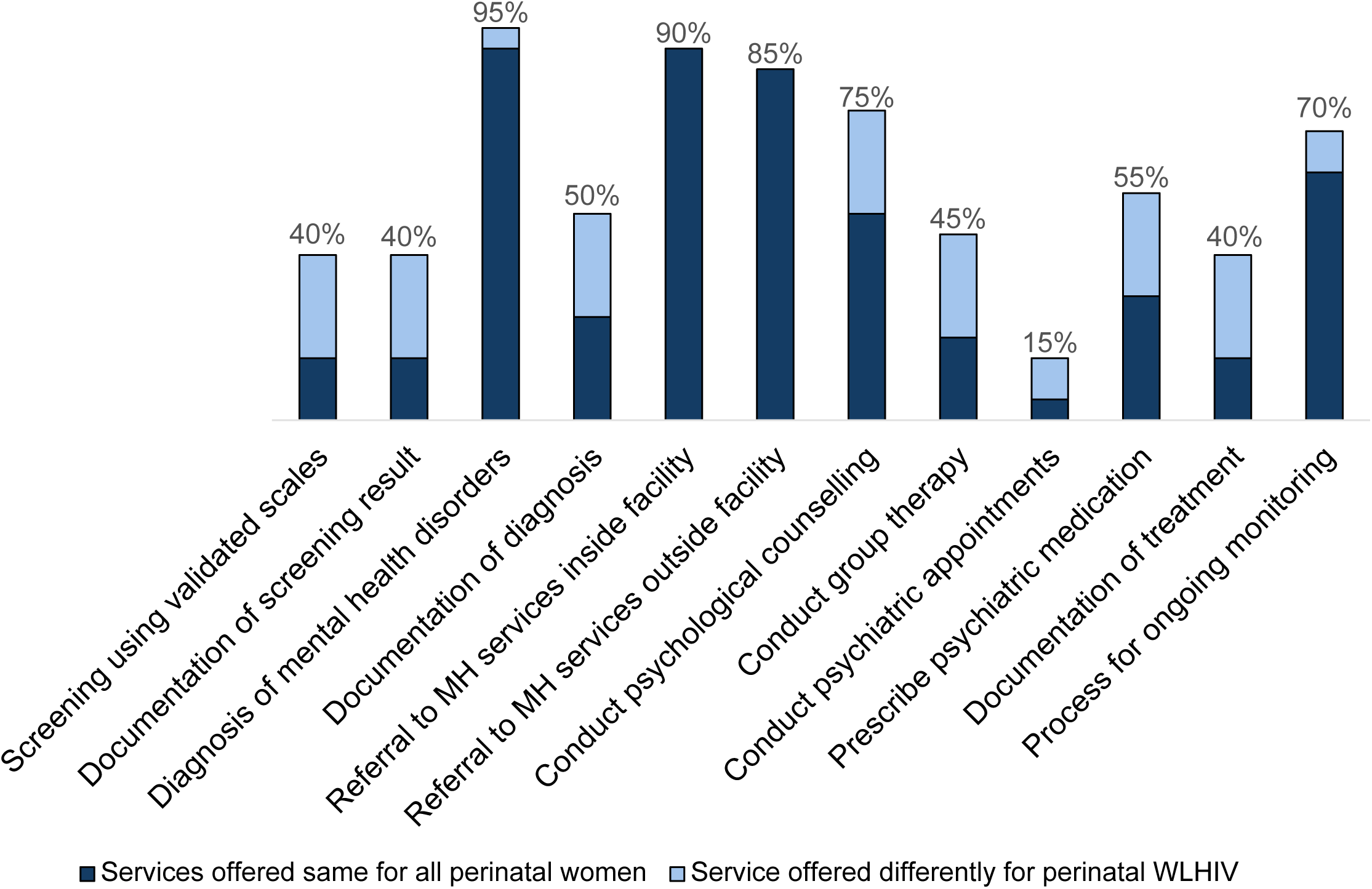
Perinatal mental health services offered in MCH facilities in Western Kenya (N=20)

Overall, 18 (90%) facilities provided linkage to perinatal mental health services within the facility and 17 (85%) provided referral outside of the facility. Nurses were the cadre most frequently offering mental health referrals within the facility (n=13/18, 72%), while referral outside the facility was mainly done by clinical officers (n=15/17, 88%). Fifteen (75%) health facilities reported offering some form of psychological counseling (either evidence-based psychological interventions or general counseling), primarily by a medical officer (n=11/15 73%) or clinical officer (n=11/15, 73%), and 9 (45%) facilities reported offering group therapy, most commonly by medical officers (n=7/9, 78%). Overall, a little over half (11, 55%) of facilities reported prescribing psychiatric medications, and three (15%) were visited monthly by a psychiatrist typically based outside of the facility. Clinical officers were the leading cadre prescribing psychiatric medication (n=9/11, 82%) and the only cadre facilitating psychiatric appointments (n=3/3, 100%) for perinatal patients with PMAD. Among facilities referring women to internal mental health services, only 44% of facilities (8/18) reported documenting perinatal mental health treatment in clinical records. Fourteen facilities (70%) reported having procedures to monitor PMAD patients over time, which was primarily conducted by clinical officers (n=9/14, 64%), followed by nurses (n=5/15, 36%). Across the perinatal mental health care cascade, five facilities consistently reported offering services differently among WLHIV compared to other perinatal women. These facilities differentiated care based on HIV status for perinatal mental health screening, documentation of screening and diagnosis results, provision of individual and group psychotherapy, and prescription of psychiatric medication and documentation.

### 3.3 Psychological intervention provision

We further explored whether evidence-based psychological therapies such as cognitive behavioral therapy (CBT), problem management plus (PM+), or interpersonal psychotherapy (IPT) were provided within MCH facilities. A high proportion of facilities initially reported providing evidence-based psychological interventions (range across evidence-based psychological therapies listed in survey: 40-70%). These services were primarily delivered by nurses, “mentor mothers,” or adherence counsellors. Less frequently, these services were delivered by psychiatrists, psychologists, or social workers. However, in response to follow-up questions, none of the facilities reported using manualized evidence-based therapies.

### 3.4 Mental health medication available in MCH facilities in Western Kenya

Figure 2 represents the availability of unexpired mental health medication in the MCH facilities in Western Kenya. Most of the sites (n=16/20, 80%) reported having at least one type of anxiolytic, most commonly diazepam (n=15/20, 75%). Over half of the facilities (n=13/20, 65%) offered antiepileptics, most commonly phenobarbitone (n=11/20, 55%). Half of the facilities (n=10/20, 50%) offered antidepressants, most commonly amitriptyline (n=10/20, 50%), and half of the facilities offered carbamazepine (n=10/20, 50%) as a mood stabilizer. Less than half of the facilities (n=9/20, 45%) reported offering antipsychotic drugs, with chlorpromazine and olanzapine being the most commonly available (n=4/20, 20% each). Overall, the majority of facilities only had first-generation medications available. Two facilities offered a second-generation antidepressant (fluoxetine) and four facilities offered a second-generation antipsychotic (olanzapine).

**Figure 2.**
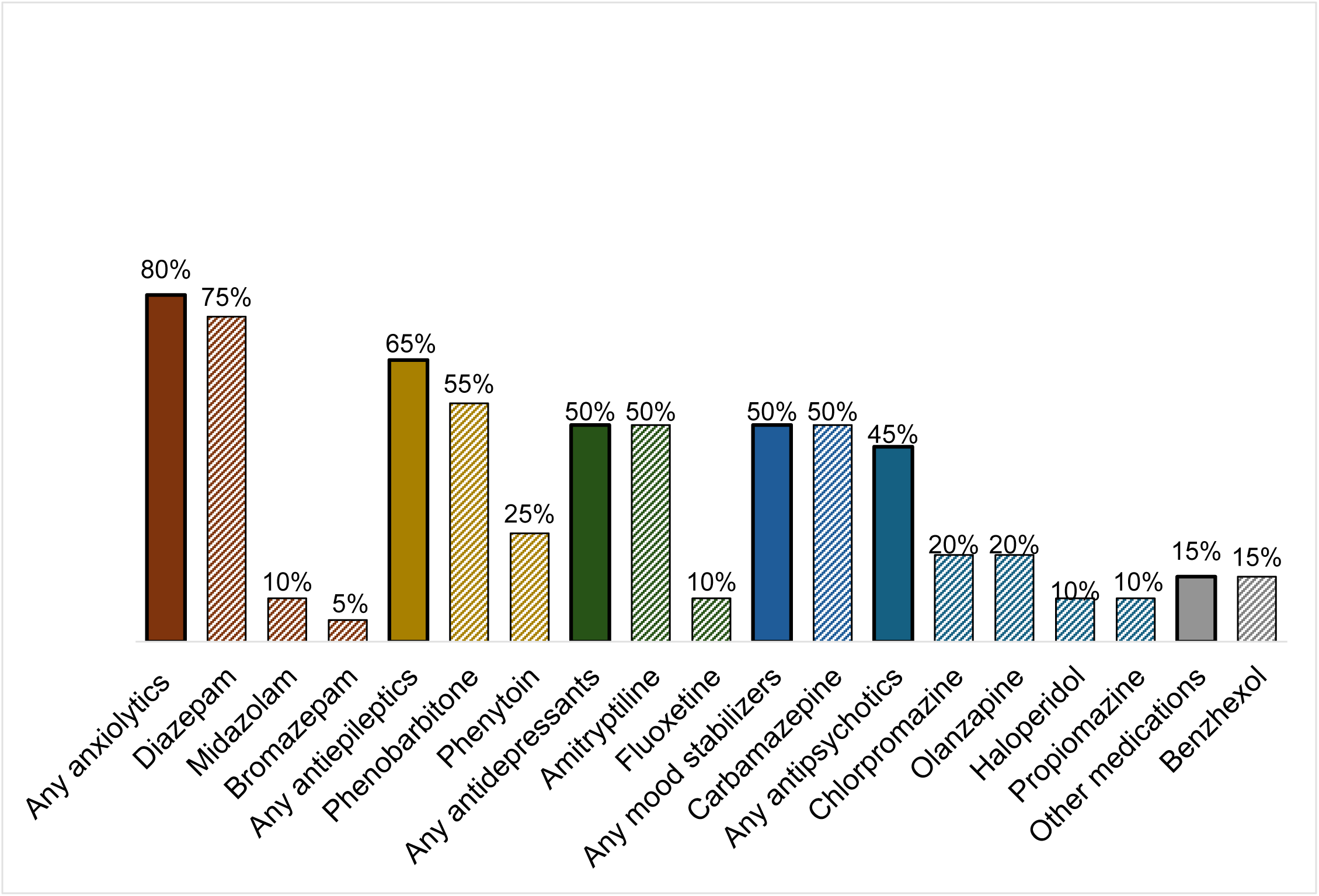
Mental health medication availability in MCH facilities in Western Kenya (N=20)

### 3.5 Infrastructure to integrate a stepped care model for perinatal mental health services

In Table 2 we summarize the infrastructure available to support the integration of the IPMH stepped-care model for perinatal mental health services (including universal screening, lay provision of PM+, and telepsychiatry.

**Table 2.** Current status to integrate a stepped care model to deliver perinatal mental health services into MCH facilities in Western Kenya.

| **Integration components** | **Western Kenya**<br>**Overall**<br>**N = 20** |
| --- | --- |
|  | **N (%) or Median**<br>**(Interquartile**<br>**range)** |
| **Screening services** |  |
| Screening for MH symptoms using a validated scale | 8 (40%) |
| **Frequency of screening conduct** |  |
| Every patient encounter | 7 (88%) |
| First encounter with patient | 1 (13%) |
| No response/No screening |  |
| **Mental health disorders screened** |  |
| Depression | 8 (40%) |
| Anxiety | 3 (15%) |
| PTSD | 1 (1%) |
| **Screening tools used** |  |
| Patient Health Questionnaire-2 | 1 (1%) |
| Patient Health Questionnaire-9 | 8 (40%) |
| Generalized Anxiety Disorder-7 | 3 (15%) |
| PTSD PCL | 1 (1%) |
| **Facility infrastructure** |  |
| Total inpatient hospital beds | 20.0 (14.0, 29.0) |
| Maternity beds | 7.5 (5.5, 13.0) |
| Mental health/psychiatric beds | 1 (5.0%) |
| Perinatal mental health/psychiatric beds | 1 (100%) |
| **Healthcare workforce** |  |
| Overall medical staff | 37.5 (31.0, 45.5) |
| Medical Officers | 0.0 (0.0, 0.0) |
| Psychiatrists | 0.0 (0.0, 0.0) |
| Obstetricians/gynecologists | 0.0 (0.0, 0.0) |
| Nurses | 9.0 (6.0, 12.5) |
| Psychologists | 0.0 (0.0, 0.0) |
| Mental health social workers | 0.0 (0.0, 0.0) |
| Clinical officers | 5.0 (3.0, 6.0) |
| HIV Testing Services counsellors | 3.0 (2.0, 3.0) |
| Community health workers | 17.0 (12.0, 23.5) |
| Mentor mothers | 2.0 (2.0, 3.0) |
| Psychiatric nurses | 0.0 (0.0, 0.0) |
| Counsellors (e.g., lay, behavioral) | 1.0 (1.0, 1.5) |
| Other mental health workers | 0.0 (0.0, 0.0) |
| **Telemedicine infrastructure** |  |
| Functioning land-line phone available | 2 (10%) |
| Functioning cellular phone available | 18 (90%) |
| Functioning computer available | 18 (90%) |
| Internet access today | 15 (75%) |
| Electricity continuously available during facility working hours (past 7 days) | 17 (85%) |
| Facility working hours per day | 20 (100%) |
| Water source available in facility premises | 20 (100%) |
| Patient consultation room with auditory/visual privacy available | 20 (100%) |
| **Medical records system** |  |
| Paper-based | 20 (100%) |
| Electronic | 19 (95%) |
| Other | 0 (0%) |
| **Mental Health Integration level** |  |
| Level 1 | 6 (30%) |
| Level 2 | 8 (40%) |
| Level 3 | 6 (30%) |
| Level 4 | 0 (0%) |
| Level 5 | 0 (0%) |

### 3.6 Screening services

Among the eight facilities offering screening services using a validated scale, most reported performing screening at every patient encounter (n=7/8, 88%) while one facility reported screening at the first patient encounter only (n=1/8, 13%). Under half of the facilities offered screening for depression, using the patient health questionnaire-9 (PHQ-9) scale (n=8/20, 40%). Under a quarter offered screening for anxiety, using the generalized anxiety disorder-7 (GAD-7) scale (n=3/20, 15%). One facility (5%) offered screening for post-traumatic stress disorder (PTSD), using the PTSD PCL-5.

### 3.7 Physical and telemedicine infrastructure

Facilities reported a median of 20.0 inpatient beds (interquartile range [IQR]: 14.0-29.0) with a median of 7.5 beds designated for maternity patients (IQR: 5.5-13.0). Only one facility (5%) reported having a dedicated bed for mental health, and one facility (5%) reported a bed dedicated to perinatal mental health. Most facilities (n=19/20, 95%) used electronic medical record systems, and all facilities also utilized paper-based medical records (n=20/20, 100%).

All 20 (100%) facilities had a patient consultation room available with audio-visual privacy which could be utilized to facilitate telemedicine visits for perinatal mental health. The majority of facilities (n=18/20, 90%) had a facility-owned cellular phone available, 2 (10%) had a landline telephone, 18 (90%) had a computer available, and 15 (75%) reported having internet available on the day of the survey. Most facilities (n=17/20, 85%) reported continuous electricity during facility working hours in the prior 7 days; all 20 facilities (100%) reported water sources available on the facility premises.

### 3.8 Healthcare workforce

The overall medical staff across the 20 facilities surveyed had a mean of 45 medical staff members (standard deviation [sd]: 19.8). Community health workers were the most common cadre with an average of about 20 community health workers per facility (mean: 19.8, sd: 10.8). Facilities had an average of 10 nurses (mean: 10.1, sd: 5.6), five clinical officers (mean: 5.1, sd: 3.0), three HIV testing services counselors (mean: 3.1, sd:1.9), and two mentor mothers (mean: 2.3, sd:1.0).

### 3.9 Mental health integration level in MCH facilities in Western Kenya

Using the SAMHSA Framework for Integration, we worked with facility managers to identify the level of mental health integration in MCH facilities. Approximately a third of facilities (n=6/20, 30%) reported minimal collaboration across separate facilities with rare communication, making them “Level 1” for integration. Nearly half (n=8/20, 40%) reported engaging in basic collaboration to provide mental health care at a distance across separate facilities with periodic collaboration (Level 2). The remaining six facilities reported basic collaboration on site within the same facility across separate systems with regular communication (Level 3). None of the facility managers felt their facility fit into levels 4 or 5 for integration of mental health services within MCH services, characterized by close collaboration within the same space and system.

## 4. DISCUSSION

In this descriptive evaluation of mental health services within MCH clinics in Western Kenya, we observed that screening and referral to mental health services are currently not offered in a structured and systematic manner, which presents a serious challenge to effectively alleviating perinatal mood and anxiety disorders. However, MCH settings have existing infrastructure, resources, and personnel that offer opportunities to support integration of perinatal mental health care into these routinely attended clinics. Our findings extend the literature about the current state of perinatal mental health services in the African region and inform efforts to expand access, including our team’s “Integrated Perinatal Mental Health Program (IPMH)”. IPMH will evaluate an evidence-based treatment model to improve perinatal mental health integrated into MCH clinics in Western Kenya.

We observed that screening for PMAD using a validated scale, and documenting results was uncommon across facilities in Western Kenya. This is consistent with findings from a study on Stepped Care for Maternal Mental Health in South Africa that revealed a lack of routine screening or treatment of maternal mental disorders in primary care settings (19). Integrating mental health screening into routine antenatal procedures in low-resource settings, where common mental disorders are often overlooked, has been found to have the potential to narrow treatment gap significantly (16,17,19). There is evidence that screening alone can have some benefits by raising awareness of one’s depressive or anxiety symptoms, although initiation of treatment or referral to mental health care providers offers the maximum benefit (20). Our evaluation identified that women living with HIV were more likely to receive PMAD screening as compared to their HIV negative counterparts in Western Kenya. Further, depression was a commonly screened mental health disorder. However, low screening rates overall observed in our survey confirm a need to implement interventions that will increase access to PMAD screening for early detection. Studies have shown that early detection of PMAD symptoms through routine screening of depression and anxiety in the obstetrics setting is key to alleviating the significant consequences of PMAD symptoms (13),

Despite gaps in validated PMAD screening, facility managers at most of the sites reported that mental health diagnoses occurred in that site. Our survey did not require facility managers to describe their procedures for mental health diagnosis; thus, we cannot discern whether PMAD diagnoses are conducted using a standardized clinical interview or less systematic provider judgment. Further, there was a gap in documenting and following up on diagnoses. Only three sites made psychiatric appointments, and less than half of the facilities documented treatment for perinatal mental health concerns. These findings echo results from a study in South Africa, which observed that despite high levels of antenatal and postnatal depression, there was no treatment for maternal mental disorders in primary care settings (19). There is strong evidence from multiple studies that psychological and psychosocial interventions for perinatal depression are effective and cost-effective (19,21). None of the sites surveyed in the present evaluation offered evidence-based psychological interventions according to the training, supervision, or guidance from published intervention manuals. However, some form of supportive talk therapy is available at a majority of MCH facilities surveyed; these talk therapies and supportive counseling approaches do not appear to be standardized, and their effectiveness has not been evaluated. There is reasonable evidence for the benefit of psychological interventions in perinatal depression and anxiety in LMICs (22). A systematic Review of Interventions That Integrate Perinatal Mental Health Care into Routine Maternal Care in Low- and Middle-Income Countries support our finding that evidence-based psychological interventions for perinatal mental health are largely unavailable (23)

Among the facilities reporting use of psychotropic medication, the majority only had first-generation treatment available (e.g., chlorpromazine, amitriptyline), which may not be well-suited for perinatal women. Only two facilities reported offering a second-generation antidepressant, and 4 facilities reported a second-generation antipsychotic that may be more appropriate for this vulnerable population. These findings parallel those of a study that indicated limited availability of essential mental health medicines in LMICs (24). Appropriate mental health medication for perinatal women, such as sertraline, is lacking within MCH facilities, potentially highlighting a gap in knowledge of and access to newer medications. Integration of evidence-based psychological therapies and expanded access to psychiatric medications in MCH settings would dramatically improve access to perinatal mental health support. As policy makers become more aware of the need for identification and treatment of PMAD, it is also important to highlight the urgent need to increase access to evidence based treatments in these settings.

Our study explored availability of infrastructure, resources, and personnel necessary to integrate a stepped care model for perinatal mental health services into MCH settings of Western Kenya, we observed that only one facility had a dedicated bed for perinatal mental health. Further, most health facilities lacked specialized mental health professionals. A systematic review of interventions for perinatal depression in low and middle-income countries similarly observed the sparse specialized human resources for mental health, with insufficient numbers of psychiatrists, psychologists, social workers, and mental health nurses available to meet the demand (22). Despite the few mental health specialists, the study observed that there were adequate healthcare providers across the sites in other cadres, primarily nurses, whose skillsets could be leveraged to support mental health service integration and delivery. Task shifting is an evidence-based approach to strengthening and expanding the health workforce whereby responsibilities are shifted from specialized health workers to health worker or peer provider cadres with lower training and/or fewer qualifications (WHO, 2008). In the context of mental health screening and treatment, task shifting represents a shift from mental health professionals to healthcare workers who lack formalized training in mental health (25). Additionally, studies have shown that technology-based approaches like telephone-delivered and computer-delivered interventions can help scale mental healthcare and support (26,27). Our study indicated that most facilities had a cellular phone available that could be used for telepsychiatry, as well as internet access, a computer, continuous electricity, and a private patient consultation room.

We worked with the facility managers to identify the existing level of mental health integration in MCH facilities using the SAMHSA Framework for Integration. Most of the sites reported minimal collaboration to engage in basic collaboration to provide mental health care at a distance. Sites with higher existing levels of integration only showed evidence of basic onsite collaboration between MCH and mental health services. Currently, none of the facilities surveyed demonstrate close collaboration between MCH and mental health or full-service integration. Globally, evidence indicates that a multilevel approach to addressing maternal mental health problems is required(28). Integrating mental health care in maternal and child health programs is an obvious next step toward ensuring equitable, accessible, and holistic mental health services are offered to pregnant and postpartum mothers(17).

### 4.1 Limitations

Our evaluation had several limitations. We collected data from a single facility manager at each of the surveyed facilities. The views of the facilities managers alone may not have been comprehensively reflective of the situation of perinatal mental health service delivery in the clinics. Survey responses were self-reported by facility managers; thus responses may be biased by recall or social desirability bias. However, a key strength of the study was the introduction of a second wave of data collection when there were uncertainties about the accuracy of reported evidence-based interventions. During this phase, the team sought physical verification through psychological intervention manuals and training materials, thereby enhancing data reliability and validity. Our study surveyed facility managers at a single time point; thus, we were not able to report changes in mental health service provision over time. Any other strengths?

## 5. CONCLUSIONS

Findings from our evaluation of perinatal mental health services within maternal child health settings in Western Kenya highlight the need to expand mental health screening, documentation of mental health diagnoses, availability of psychiatric medication, and evidence-based psychological interventions. Maternal child health settings within Kenya currently have human resources and infrastructure available to support integration of a stepped care model for perinatal mental health services, which could include task-shifted mental health screening, provision of psychological therapy, and tele-linkage to specialist mental health services, as needed. Our results highlight specific, actionable areas to address toward achieving the Kenya Mental Health Action Plan 2021-2025, which advocates for expanding access to mental health services via existing and well-attended maternal child health and HIV care programs. Future initiatives to support perinatal mental health in Kenya should leverage the existing infrastructure and resources within these care settings while systematically expanding screening, diagnosis, and treatment approaches to improve perinatal mental health.

## Data Availability

Data are available upon reasonable request

## Notes

### Competing Interest Statement

The authors have declared no competing interest.

### Funding Statement

The authors have no financial conflicts of interest to declare. This study was funded through US National Institutes of Mental Health (R01MH133266) The funding agency had no role in writing this manuscript or submitting it for publication.

### Author Declarations

This study was approved by the ethical review committees both at Kenyatta National Hospital / University of Nairobi (ERC number P425/04/2023) and at the University of Washington (IRB number 00017933).

